# Genome-Wide Association Study Meta-Analysis Uncovers Novel Genetic Variants Associated with Olfactory Dysfunction

**DOI:** 10.1101/2024.08.09.24311665

**Authors:** Mohammed Aslam Imtiaz, Konstantinos Melas, Adrienne Tin, Valentina Talevi, Honglei Chen, Myriam Fornage, Srishti Shrestha, Martin Gögele, David Emmert, Cristian Pattaro, Peter Pramstaller, Franz Förster, Katrin Horn, Thomas H. Mosley, Christian Fuchsberger, Markus Scholz, Monique M.B. Breteler, N. Ahmad Aziz

## Abstract

**IMPORTANCE:** Olfactory dysfunction is among the earliest signs of many age-related neurodegenerative diseases and has been associated with increased mortality in older adults; however, its genetic basis remains largely unknown.

**OBJECTIVE:** To identify the genetic loci associated with olfactory dysfunction in the general population.

**DESIGN, SETTING AND PARTIICIPANTS:** This genome-wide association study meta-analysis (GWMA) included participants of European ancestry (N = 22,730) enrolled in four different large population-based studies, followed by a multi-ancestry GWMA including participants of African ancestry (N = 1,030). The data analysis was performed from March 2023 through June 2024.

**EXPOSURES:** Genome-wide single nucleotide polymorphisms.

**MAIN OUTCOMES AND MEASURES:** Olfactory dysfunction was the outcome and assessed using a 12-item smell identification test.

**RESULTS:** GWMA revealed a novel genome-wide significant locus (tagged by rs11228623 at 11q12) associated with olfactory dysfunction. Gene-based analysis revealed a high enrichment for olfactory receptor genes in this region. Phenome-wide association studies demonstrated associations between genetic variants related to olfactory dysfunction and blood cell counts, kidney function, skeletal muscle mass, cholesterol levels and cardiovascular disease. Using individual-level data, we also confirmed and quantified the strength of these associations on a phenotypic level. Moreover, employing two-sample Mendelian Randomization analyses, we found evidence for causal associations between olfactory dysfunction and these phenotypes.

**CONCLUSIONS:** These findings provide novel insights into the genetic architecture of the sense of smell and highlight its importance for many aspects of human health.

**Key Points:** 

**Question:** What is the genetic basis of olfactory dysfunction, and is it causally related to adverse health outcomes?

**Findings:** This genome-wide association study meta-analysis (GWMA) of 22,730 European and 1,030 African participants identified a novel genomic locus, enriched for olfactory receptor genes, robustly associated with olfactory dysfunction. Two-sample Mendelian Randomization analyses provided evidence for causal associations of olfactory dysfunction with biochemical, anthropometric and cardiovascular health outcomes.

**Meaning:** These findings provide new insights into the genetic architecture of olfaction and implicate olfactory dysfunction as a causal risk factor for many aspects of human health.

## Introduction

Olfactory function is paramount to both safety and quality of life, enabling detection of hazardous or unpleasant odors, and contributing to the enjoyment of scents, food and drink. Impairment of olfaction is very common, affecting approximately 1 in 5 adults, with an increased prevalence among older individuals.^1^ Indeed, aging is a major determinant of olfactory dysfunction and is thought to affect both the central and peripheral olfactory system. Other risk factors associated with smell loss in adults include sinonasal diseases, smoking, and alcohol consumption.^1^ Moreover, reduced sense of smell is a well-established consequence of coronavirus disease 2019 and one of the earliest markers of many neurodegenerative diseases. ^2–4^ Importantly, olfactory dysfunction itself has been suggested as a risk factor associated with cognitive decline,^5^ frailty,^6^ cardiovascular diseases,^7^ kidney function,^8^ and increased mortality.^9^ However, the causality of these associations remains to be elucidated. Thus, uncovering the genetic architecture of olfactory dysfunction could not only provide novel molecular targets for its treatment, but could also be instrumental to assessing whether decreased sense of smell is causally related to adverse health outcomes.

Despite the high prevalence of olfactory dysfunction and its involvement in a variety of diseases, the genetic architecture of olfactory dysfunction remains largely unknown. Odor identification, the most commonly studied component of olfactory function, has been shown to have a low to moderate heritability.^10,11^ A previous genome-wide association study (GWAS) of olfactory function identified nine genome-wide significant loci associated with odor identification among African Americans (N = 1,979), but only two among European Americans (N = 6,582).^12,13^ Interestingly, many of these regions were related to neurodegenerative and neuropsychiatric diseases.^12,13^ More recently, Raj et al. examined the association between single nucleotide polymorphisms (SNPs) located in or near olfactory receptor genes (32,282 SNPs) and the ability to identify individual odors, detecting a larger number of SNPs (9,267 SNPs) at a suggestive statistical significance level (p < 0.001). However, none of these SNPs remained significant after adjustment for multiple testing, failing to replicate the findings from the previous GWAS.^14^

We aimed to further elucidate the genetic architecture of olfactory dysfunction by performing the largest GWAS and meta-analysis of sense of smell to date, among adults of European and African ancestry, using data from four different large-scale, population-based studies. Moreover, using a two-sample Mendelian Randomization (MR) approach, we investigated the causal relationship between olfactory dysfunction and different health-related outcomes.

## Methods

### Study population

We included 1,030 individuals of African American ancestry (AAs) from the ARIC Study,^15^ and 22,730 individuals of European ancestry (EUR) from the Rhineland Study, the ARIC Study, the LIFE-Adult-Study^16^ and the Cooperative Health Research in South Tyrol (CHRIS) study,^17^ who had complete genetic and olfactory function data (**Table 1**).

**Table 1.** Baseline characteristics of participating cohorts.

| Characteristic | RHINELAND | ARIC<br>EA | ARIC<br>AA | LIFE-ADULT | CHRIS |
| --- | --- | --- | --- | --- | --- |
| N | 6580 | 3654 | 1030 | 4771 | 6696 |
| Age (years) | 55.9 ± 13.5 | 75.9 ± 5.2 | 75.0 ± 5.1 | 57.4 ±<br>12.97 | 44.6 ±<br>16.5 |
| Women, % | 55.4 | 56.0 | 66.0 | 51.4 | 53.2 |
| Olfactory<br>dysfunction score<br>(mean ± SD) | 2.10 ± 1.71 | 2.48 ± 2.29 | 4.01 ± 2.59 | 2.06 ±<br>1.72 | 1.6 ± 1.55 |
| *APOE* *e4 allele<br>carriers, % | 1233 (26%) | 922 (25%) | 394 (38%) | 1148<br>(24.1%) | 2396<br>(35%) |
| Global cognition<br>score<br>(mean ± SD) | -0.57 ± 0.55 | - | - |  |  |
| MMSE<br>(mean ± SD) | - | 27.9 ± 2.3 | 25.6 ± 3.3 |  | 28.98 ±<br>1.62 |
| Cognition<br>(CERAD) |  |  |  | 24 ± 6.34 |  |

**Abbreviations:** AA: African-American; ARIC: Atherosclerosis Risk in Community Study; CHRIS: The Cooperative Health Research in South Tyrol; EA: European-American; EUR: European; LIFE-Adult: LIFE-Adult Cohort; MMSE: Mini-Mental State Examination, range of possible score 0-30; CERAD: Consortium to Establish a Registry for Alzheimer's disease Neuropsychological Battery; RHINELAND: Rhineland Study; SD: Standard deviation.

### Assessment of olfactory function

In the Rhineland Study, the ARIC Study, and the LIFE-Adult-Study, olfactory function was assessed using the 12-item "Sniffin’ Sticks" odor identification test (SIT-12), a widely utilized screening instrument for assessing odor identification ability.^18^ In the CHRIS study, the 16-item "Sniffin’ Sticks" odor identification test was employed and for this analysis restricted to the SIT-12 items. Olfactory dysfunction was defined as the total number of incorrectly identified odors on the SIT-12 test (range 0-12) (**eMethods** in **Supplement 1**).

### Genotyping, quality control and imputation

Genotyping was performed in all four cohorts using commercially available genetic arrays (**eMethods** in **Supplement 1**), followed by standard quality control measures. In brief, quality control was performed using PLINK (version 1.9), excluding SNPs based on poor genotyping rate (< 99%), minor allele frequency (MAF) < 1% or Hardy-Weinberg Disequilibrium (p < 1 x 10^-6). Imputation of genotypes was performed through IMPUTE (version 2),^19^ using as reference panels 1000 Genomes phase 3 version 5 in the Rhineland Study and the LIFE-Adult cohort, 1000 Genomes version 1 phase 3 in the ARIC Study, and TOPMed in the CHRIS cohort.^20^ Variants with imputation quality score below 0.3 were excluded.^21^

### Genome-wide association studies

We performed an ancestry-specific GWAS of olfactory dysfunction in each cohort separately, using Generalized Linear Mixed Model Association Tests (GMMAT).^22^ Since olfactory dysfunction was defined as a count variable and followed a Poisson distribution, we applied a log-link function to model the association of each SNP with olfactory dysfunction, using the score test for computational efficiency.^22^ For variants that were genome-wide significant (p < 5 x 10^-8) based on the score test, we rerun the analyses while applying the Wald test to obtain estimates for the effect sizes and associated standard errors. In model 1, we adjusted for age, sex and the first 10 genetic principal components to account for population structure. In model 2, we additionally adjusted for APOE*e4 carrier/non-carrier status and global cognitive function as these factors have previously been associated with a poor sense of smell.^12^

### Meta-analysis of genome-wide association studies

The score test- and the Wald test-based results from GMMAT were meta-analyzed using the sample size-weighted or the fixed effects inverse variance-weighted method, respectively, as implemented in the meta-analysis tool for genome-wide association scans (METAL).^23^ Additionally, we performed a multi-ancestry meta-analysis by combining GWAS results from both European and African ancestry participants using a random effects model as implemented in METASOFT.^24^ The genome-wide statistical significance threshold was set at p < 5 x 10^-8.

### Genomic risk loci

We used the Functional Mapping and Analysis of GWAS (FUMA) platform to identify genomic risk loci.^25^ Genome-wide significant SNPs in relatively high linkage disequilibrium (LD) (i.e., r^2^ ≥L 0.6) with nearby SNPs were used to define genomic risk loci, merging LD blocks of independently significant SNPs within 250 kb of each other into a single genomic locus. Within each genomic locus, we defined the *lead* SNPs as those SNPs that are independent of one another at r^2^ < 0.1, using the 1000 Genome Phase 3 reference panel. **(e Methods in Supplement 1)**.

### Phenome-wide association studies and confirmation of phenotypic associations using individual-level data

We used the Open Target Genetics platform^26^ to perform phenome-wide association studies (PheWAS) for the systematic identification of phenotypes associated with genetic variations related to olfactory dysfunction. The Benjamini-Hochberg false discovery rate (FDR) method was used for multiple comparisons adjustment. In addition, using individual-level data from the Rhineland Study, we assessed whether phenotypes associated with genetic variants of olfactory dysfunction (after FDR correction), were also associated with a poor sense of smell on a phenotypic level. To this end, we employed multivariable regression models with statistical significance inferred at FDR-adjusted p < 0.05 (**eMethods** in **Supplement 1**).

### Two-sample Mendelian Randomization

We employed a two-sample Mendelian Randomization (MR) approach to test whether the associations between olfactory dysfunction and the phenotypes identified in the previous step were causal, using the TwoSample MR package.^27^ For the outcomes, we obtained GWAS summary statistics using the IEU GWAS database (**eTable13** in **Supplement 2**).^27^ To assess the risk of weak instrument bias, we calculated the F-statistic for the selected genetic instruments (**eMethods in Supplement 1**).^28^

## Results

### Population characteristics

The study and ancestry specific population characteristics are provided in **Table 1**. On average, participants of European ancestry had a lower degree of olfactory dysfunction and scored higher on cognitive tests compared to those of African ancestry. Moreover, APOE*e4 allele carrier frequency was lower in participants of European ancestry.

### GWAS meta-analysis

The GWAS meta-analysis of European ancestry participants identified 22 and 1523 genome-wide significant SNPs located on chromosome 11 based on results from model 1 and model 2, respectively (**Figure 1**). Overall, the genomic inflation factor (λ) in each European cohort was low, ranging from 0.49 t to 1.01 (**eFigure 1 in Supplement 1**). Because the λ-value was relatively low (0.49) in the ARIC European ancestry cohort, indicating genomic deflation, we also performed a sensitivity analysis in which we corrected the p-values in this group by dividing the chi-squared statistic by λ,^29^ and re-running the European-based meta-analysis. This, however, did not change the results (**eFigure 2 in. Supplement 1**). In model 2, the meta-analysis identified one lead SNP (rs11228623), as well as three independent significant SNPs (rs12786376, rs34099256, and rs369532258), across one genomic risk locus (11q12) (**Table 2**, **Figure 2**, and **eTables 1-4 in Supplement 2**). In the multi-ancestry GWAS meta-analysis, the associations of the lead SNP (rs11228623) and one of the independent SNPs (rs12786376) with olfactory dysfunction remained directionally consistent and genome-wide significant (**Table 2**).

**Figure 1.** Genome-wide association meta-analysis of olfactory function. Manhattan (**A**) and corresponding quantile-quantile plot (**B**) of the genome-wide meta-analysis of sense of smell in people of European ancestry for model 1. Manhattan (**C**) and corresponding quantile-quantile plot (**D**) of the genome-wide meta-analysis of the sense of smell in people of European ancestry for model 2. The horizontal red dashed lines indicate the threshold for genome-wide significance (i.e., p < 5 x 10^-8).

**Figure 2.** Regional plots of the lead and candidate genetic variants. The figure shows positional mapping of the 11q12 locus with the top lead single nucleotide polymorphism (SNP), as well as variants in linkage disequilibrium with this SNP according to the r^2^-color coded key.

**Table 2.**
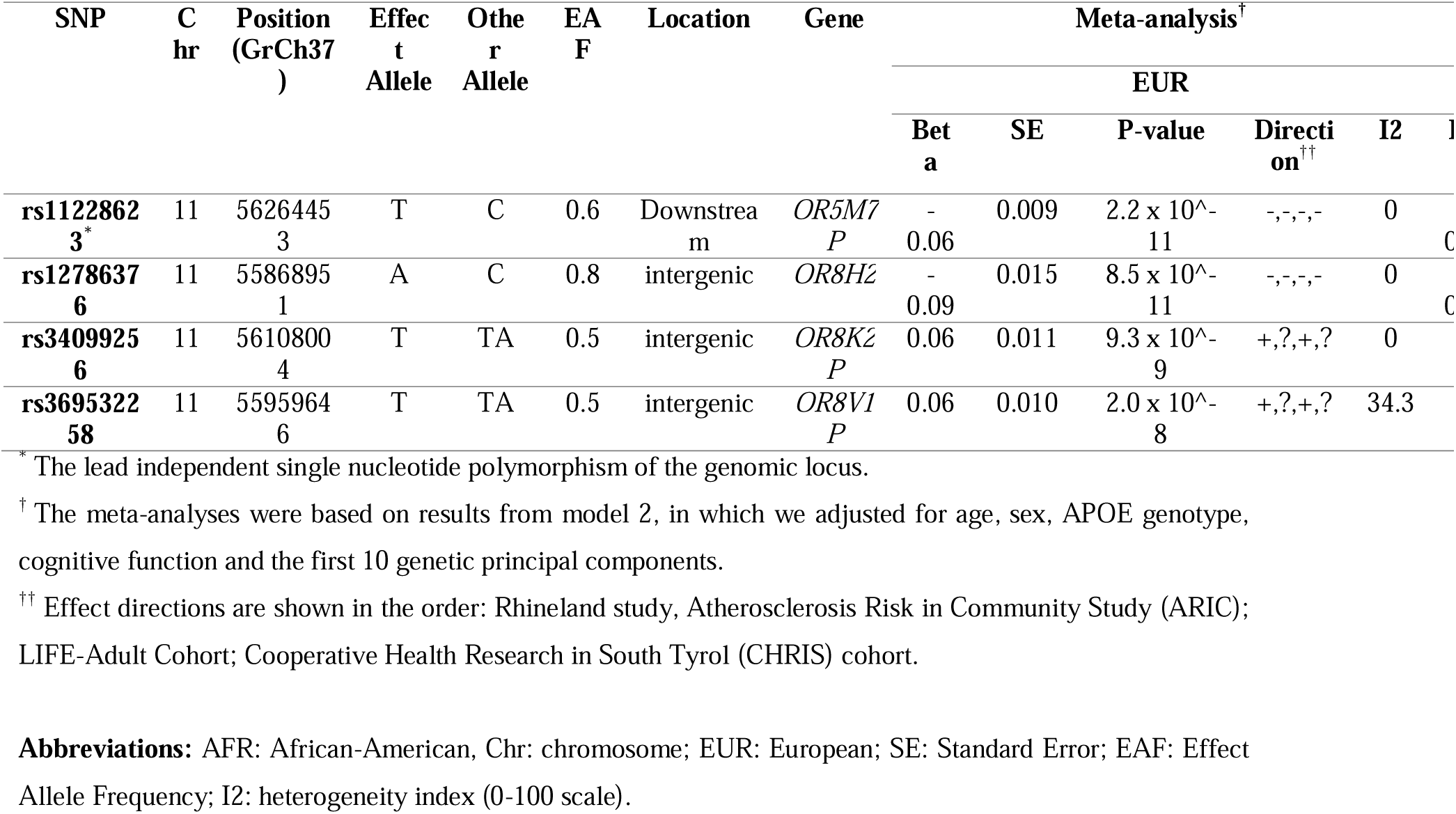
Independent genome-wide significant single nucleotide polymorphisms (r^2^ < 0.6 and p < 5 x 10^-8) associated with olfactory dysfunction in European GWAS meta-analysis, and comparison with the cross-ancestry meta-analysis. The results displayed are summary statistics derived from the GWAS meta-analysis of Wald test results.

The olfactory dysfunction-associated SNPs were mapped to genes based on positional, eQTL and chromatin interaction mapping. Positional mapping identified 34 olfactory receptor genes. We discovered 3 genes (*OR5M11, SLC43A3 and PRG2*) based on eQTL mapping of which one gene (*OR5M11)* overlapped with those identified through positional mapping (**eTables 5** & **6 in Supplement 2**). Chromatin interaction mapping, based on Hi-C data, showed significant (FDR < 1 x 10^-6) chromatin interactions between enhancers of candidate genes in this region and the promoter regions of *MPEG1, LRR45,* and *OR4A16,* as well as those of several other genes on chromosome 11q12 (**eFigure 3** in **Supplement 1** & **eTable7** in **Supplement 2**). After Bonferroni-correction, gene-based analysis using MAGMA identified 21 genome-wide significant (p < 2.6 x 10^-6) genes, with *OR5M11* as the top hit (p < 8.4 x 10^-9) (**eTable8** in **Supplement 2**). In the MAGMA gene-set enrichment analysis, the top gene sets were enriched for “reactome hedgehog ligand biosynthesis” and “reactome degradation of beta catenin by the destruction complex” (**eTable9** in **Supplement 2**), but none survived multiple testing correction. Interrogation of MSigDB showed that the mapped genes at the 11q12 locus were significantly enriched for pathways related to “general odorant binding proteins, sensory perception of smell”, “molecular function, odorant binding” and “Grueneberg Ganglion, olfactory transduction” (**eFigure 4** in **Supplement 1**).

**Figure 3.** Comparison of phenotype-level and Mendelian Randomization estimates for associations between olfactory dysfunction and different traits and diseases identified through phenome-wide association studies. **A)** Forest plot depicting associations between olfactory dysfunction and other phenotypes (identified through phenome-wide association studies after false discovery rate correction) using individual-level data from the Rhineland Study. The standardized regression estimate indicates the change in standard deviations in the outcome for one standard deviation increase in olfactory dysfunction. **B)** Forest plot showing causal estimates from two-sample Mendelian Randomization analyses of the effect of olfactory dysfunction on other phenotypes (Wald ratio test). The regression estimate indicates the change in standard deviations in the outcome for the effect allele of the lead genetic variant (for binary outcomes, including hypertension, heart failure, coronary artery diseases, the regression estimate refers to the logarithm of the odds ratio).

### Gene expression analysis

Of the 41 genes identified through positional, eQTL and chromatin interaction mapping, expression levels were available for *MPEG1* (tagged by rs12786376) and *SLC43A3* (tagged by rs1811871001) in whole blood for 1985 participants in the Rhineland Study. Olfactory dysfunction was not associated with the expression levels of these two genes. However, a borderline significant interaction with age was found for *SLC43A3*. Age-stratified analysis showed that higher *SLC43A3* expression was associated with worse olfactory function at borderline significance for participants aged 30-50 years (**eTable10** in **Supplement 2**). However, we found no significant association between rs1811871001 and *SLC43A3* expression, or rs12786376 and *MPEG1* expression.

### Phenome-wide association studies and phenotypic associations

The lead SNP (rs11228623), located within 5 kb downstream of the *OR5M7P* gene, was associated with 71 phenotypes after FDR correction (**eTable 11** in **Supplement 2**). The identified traits included lymphocyte counts, eosinophil counts, lymphocyte percentage (%) of white cells, eosinophil percentage (%) of white cells, coffee intake, pulse rate, hypertension, mean appendicular mass and levels of cystatin-C (a marker of kidney function). In participants of the Rhineland Study, we could confirm that olfactory dysfunction was indeed significantly associated with lymphocyte, neutrophil and basophil cell counts, lymphocyte percentage of white blood cells, total white blood cell counts, coffee intake, skeletal muscle mass, hand grip strength, levels of cystatin-C, heart rate, hypertension, and at borderline significance with heart failure (p < 0.07) (**Figure 3A**, and **eTable12** in **Supplement 2**).

### Two-sample Mendelian Randomization

After LD clumping, we identified one robust genetic instrument for olfactory dysfunction (rs11228623) with an F-statistic of 45.33, indicating a low probability of weak instrument bias. Two-sample MR analyses indicated causal associations between olfactory dysfunction and lymphocyte cell counts, as well as lymphocyte, neutrophil and eosinophil percentages of white blood cells, total white blood cell counts, appendicular lean mass, hand grip strength, coffee intake, hypertension, pulse rate and cardiovascular disease (**Figure 3B**, and **eTable13** in **Supplement 2**).

## Discussion

We performed the largest genome-wide meta-analysis of olfactory dysfunction to date (N=22,730), discovering 1524 genome-wide significant variants and 21 genes associated with olfactory dysfunction in people of European descent. Importantly, the novel lead SNP (rs11228623-T at 11q12) was genome-wide significant and exhibited directionally consistent effects in both ancestry-stratified and multi-ancestry analyses. Gene mapping and gene set analysis prioritized multiple genes and pathways involved in odour reception and signalling. Importantly, combining PheWAS with individual-level and MR analyses, we found evidence for a causal association between olfactory dysfunction and several anthropometric, metabolic, cardiovascular, renal and inflammatory phenotypes.

We identified a genomic risk locus for olfactory dysfunction at 11q12, enriched for olfactory receptor genes related to sensory perception of smell and olfactory transduction. The lead SNP (rs11228623-T) at this region is located downstream of the *OR5M7P* pseudogene. Using eQTL analyses we mapped the independent SNP (rs12786376-A) to three other genes (*OR5M11, PRG2* and *SLC43A3)*. Individual-level blood expression data were available for *SLC43A3*, and indicated an age-dependent association between *SLC43A3* expression levels and olfactory dysfunction. *SLC43A3* encodes a membrane transporter protein, and has been shown to control free fatty acid flux in adipocytes.^30^ To our knowledge, this is the first time this gene and its expression have been linked to olfactory dysfunction. The majority of the other identified genes are mainly expressed in the olfactory epithelium and, therefore, could not be detected in the blood transcriptome.

The results of our MR analyses indicate that olfactory dysfunction affects anthropometric, metabolic, cardiovascular, renal and inflammatory phenotypes, highlighting its detrimental effects across different organs and tissues. This included associations of olfactory dysfunction with skeletal muscle mass and hand grip strength, which have been identified before.^31,32^ A potential explanation could be that smell loss leads to changes in dietary habits, resulting in changes of muscle composition and strength. Conversely, it has been hypothesized that lifestyle factors, like exercise, or comorbidities might concurrently affect muscle strength and the neuronal determinants of olfaction.^31^ Our MR analyses support the former rather than the latter hypothesis. This is further supported by the causal association of olfactory dysfunction with coffee intake, a dietary habit, and cholesterol levels, which are dependent on diet. Similarly, we found that olfactory dysfunction was causally associated with hypertension, increased heart rate and a higher prevalence of heart failure. This could indicate that olfaction affects cardiovascular risk through dietary patterns and obesity, while brain vascular damage or even cardiovascular medication may affect olfaction.^7,33^ Olfactory dysfunction was also causally associated with white blood cell counts and percentages, particularly those of neutrophiles and lymphocytes. As with anthropometric and cardiovascular phenotypes, this association could be mediated by dietary and/or metabolic changes. Alternatively, a neuro-immune interaction may be involved, since neurotransmitter release following olfactory stimuli might modulate the immune response to enhance defence against infections, for example when pathogens are detected by the olfactory receptors.^34^ Perturbations of this neuro-immune cross-talk due to olfactory dysfunction may lead to changes in lymphocyte and neutrophil production.

## Limitations

The main limitation of our study is the relatively small number of participants from non-European ancestry; however, to the best of our knowledge, other large-scale population-based studies assessing olfactory dysfunction are currently lacking, precluding substantial increases of sample size in the near future. Moreover, we could replicate the association between the top genetic variant and olfactory dysfunction in people of European descent in those of African-American ancestry, but generalizability to other ethnic populations needs further investigation. Although in our MR analyses, we used a single SNP as an instrumental variable, the risk of horizontal pleiotropy is likely to be relatively low given the location of this variant in a region enriched for olfactory receptor genes. This was further supported by a high F-statistic for this variant, indicating a strong association between the genetic instrument and olfactory dysfunction, and thus low risk of weak instrument bias.

## Conclusions

We performed a multi-ancestry genome-wide meta-analysis of olfactory dysfunction in 22,730 individuals and found one genomic locus (11q12) robustly associated with olfactory dysfunction. Moreover, our analysis uncovered several genes such as *OR5M7P* and *OR5M11* related to olfactory dysfunction. Importantly, we demonstrate that olfactory dysfunction is causally associated with muscle strength and mass, cardiovascular diseases, cholesterol levels, kidney function and white blood cell counts and composition. Thus, our findings provide new insights into the genetic architecture of olfaction and implicate olfactory dysfunction as a causal risk factor for anthropometric, metabolic, cardiovascular, renal and inflammatory phenotypes. Given the high prevalence of olfactory dysfunction among aging populations, the genetic variants and molecular pathways identified here could facilitate development of novel preventive and therapeutic strategies against a range of different age-associated diseases.

## Data Availability

The Rhineland Study data on which this manuscript was based, are not publicly available due to data protection regulations. Access to data can be provided to scientists in accordance with the Rhineland Study Data Use and Access Policy. Data of LIFE-Adult are available in the framework of project agreements based on written requests. CHRIS data can be requested for research purposes by submitting a dedicated request to the CHRIS Access Committee.

## Author Contributions

**Mohammed Aslam Imtiaz:** Conceptualization, Methodology, Formal Analysis, Writing – Original Draft Preparation, Visualization; **Konstantinos Melas:** Methodology, Formal Analysis, Writing – Original Draft Preparation; **Adrienne Tien:** Methodology, Formal Analysis, Writing – Reviewing and Editing; **Valentina Talevi:** Methodology, Formal Analysis, Writing – Reviewing and Editing; **Honglei Chen:** Methodology, Formal Analysis, Writing – Reviewing and Editing; **Myriam Fornage:** Methodology, Formal Analysis, Writing – Reviewing and Editing; **Srishti Shrestha:** Methodology, Formal Analysis, Writing – Reviewing and Editing; **David Emmert**: Methodology, Formal Analysis, Writing – Reviewing and Editing; **Cristian Pattaro**: Methodology, Writing – Reviewing and Editing; **Peter Pramstaller**: Methodology, Writing – Reviewing and Editing; **Christian Fuchsberger:** Methodology, Formal Analysis, Writing – Reviewing and Editing; **Franz Förster:** Methodology, Formal Analysis of LIFE-Adult, Writing – Reviewing and Editing; **Katrin Horn:** Methodology, Formal Analysis of LIFE-Adult, Writing – Reviewing and Editing; **Thomas H. Mosley:** Methodology, Formal Analysis, Writing – Reviewing and Editing; **Martin Gögele**: Methodology, Formal Analysis, Writing – Reviewing and Editing; **Markus Scholz:** Methodology, Writing – Reviewing and Editing; **Monique M.B. Breteler:** Conceptualization, Methodology, Resources, Writing – Reviewing and Editing, Data Curation, Funding Acquisition, Supervision; **N. Ahmad Aziz:** Conceptualization, Methodology, Writing – Reviewing and Editing, Supervision.

## Acknowledgements

The authors thank the staff and participants of all the participating cohort studies for their important contributions. The Rhineland Study is supported by DZNE core funding at the German Center for Neurodegenerative Diseases (DZNE). This work was further supported by the Federal Ministry of Education and Research (BMBF) as part of the Diet-Body-Brain Competence Cluster in Nutrition Research (grant number 01EA1410C) and in the framework of "PreBeDem - Mit Prävention und Behandlung gegen Demenz" (FKZ :01KX2230). N.A. Aziz is partly supported by an Alzheimer’s Association Research Grant (Award Number: AARG-19-616534) and a European Research Council Starting Grant (Number: 101041677). The Atherosclerosis Risk in Communities study has been funded in whole or in part with Federal funds from the National Heart, Lung, and Blood Institute, National Institutes of Health, Department of Health and Human Services, under Contract nos. (75N92022D00001, 75N92022D00002, 75N92022D00003, 75N92022D00004, 75N92022D00005). Funding was also supported by R01HL087641 and R01HL086694; National Human Genome Research Institute contract U01HG004402; and National Institutes of Health contract HHSN268200625226C. Infrastructure was partly supported by Grant Number UL1RR025005, a component of the National Institutes of Health and NIH Roadmap for Medical Research. The authors thank the staff and participants of the ARIC study for their important contributions.

LIFE-Adult is funded by the Leipzig Research Center for Civilization Diseases (LIFE). LIFE is an organizational unit affiliated to the Medical Faculty of the University of Leipzig. LIFE is funded by means of the European Union, by the European Regional Development Fund (ERDF) and by funds of the Free State of Saxony within the framework of the excellence initiative. The CHRIS study is conducted in collaboration between the Eurac Research Institute for Biomedicine and the Healthcare System of the Autonomous Province of Bolzano-South Tyrol. Investigators thank all study participants and general practitioners of the Vinschgau/Val Venosta district, and Schlanders/Silandro and Autonomous Province of Bolzano-South Tyrol Healthcare System staff for their support and collaboration. They also Biomedicine colleagues who contributed to the study. Extensive acknowledgement is reported at <u>CHRIS acknowledgements</u> - Eurac Research. CHRIS Bioresource Research Impact Factor (BRIF) code: BRIF6107. The CHRIS Study was funded by the Autonomous Province of Bolzano-South Tyrol - Department of Innovation, Research, University and Museums and supported by the European Regional Development Fund (FESR1157).

## Data sharing statement

The Rhineland Study’s data on which this manuscript was based, are not publicly available due to data protection regulations. Access to data can be provided to scientists in accordance with the Rhineland Study’s Data Use and Access Policy. Requests for additional information and/or access to the datasets can be send to.

Data of LIFE-Adult are available in the framework of project agreements based on written requests. CHRIS data can be requested for research purposes by submitting a dedicated request to the CHRIS Access Committee.

All authors had full access to the data and take responsibility for the integrity of the data as well as the accuracy of the analysis.

---

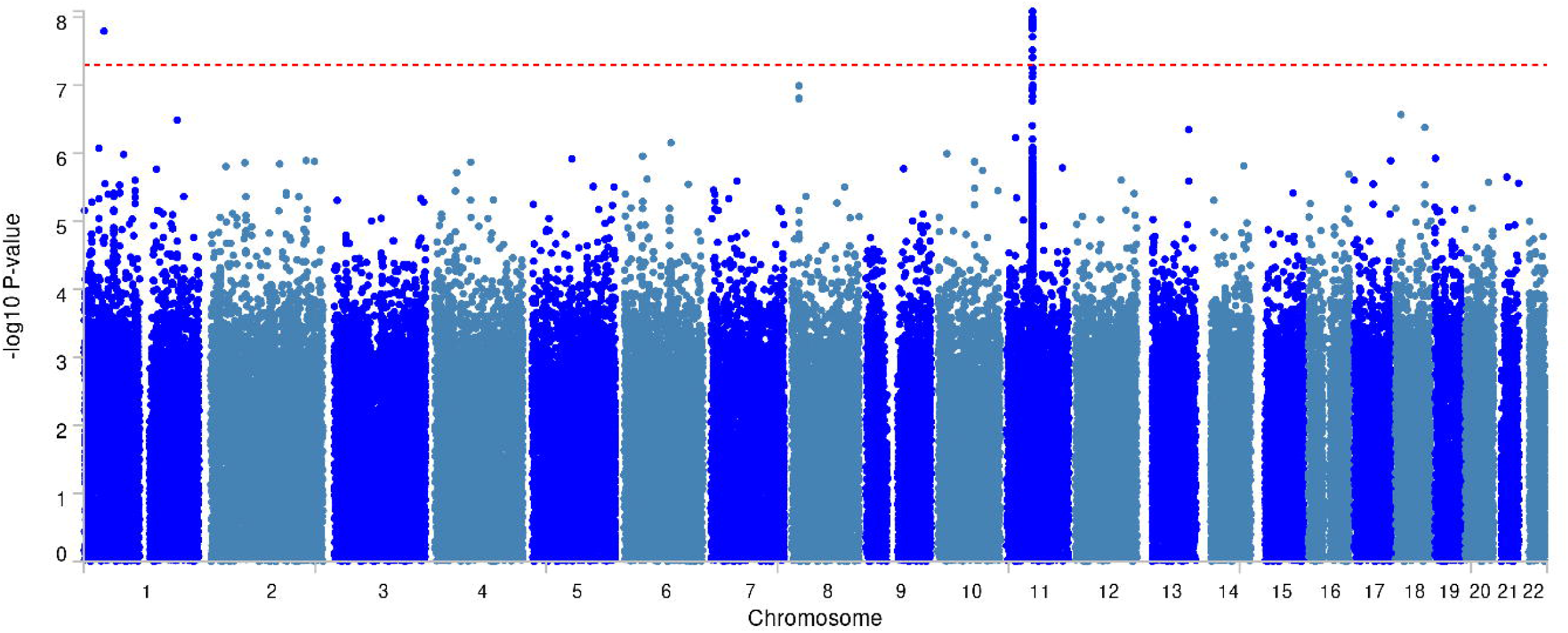

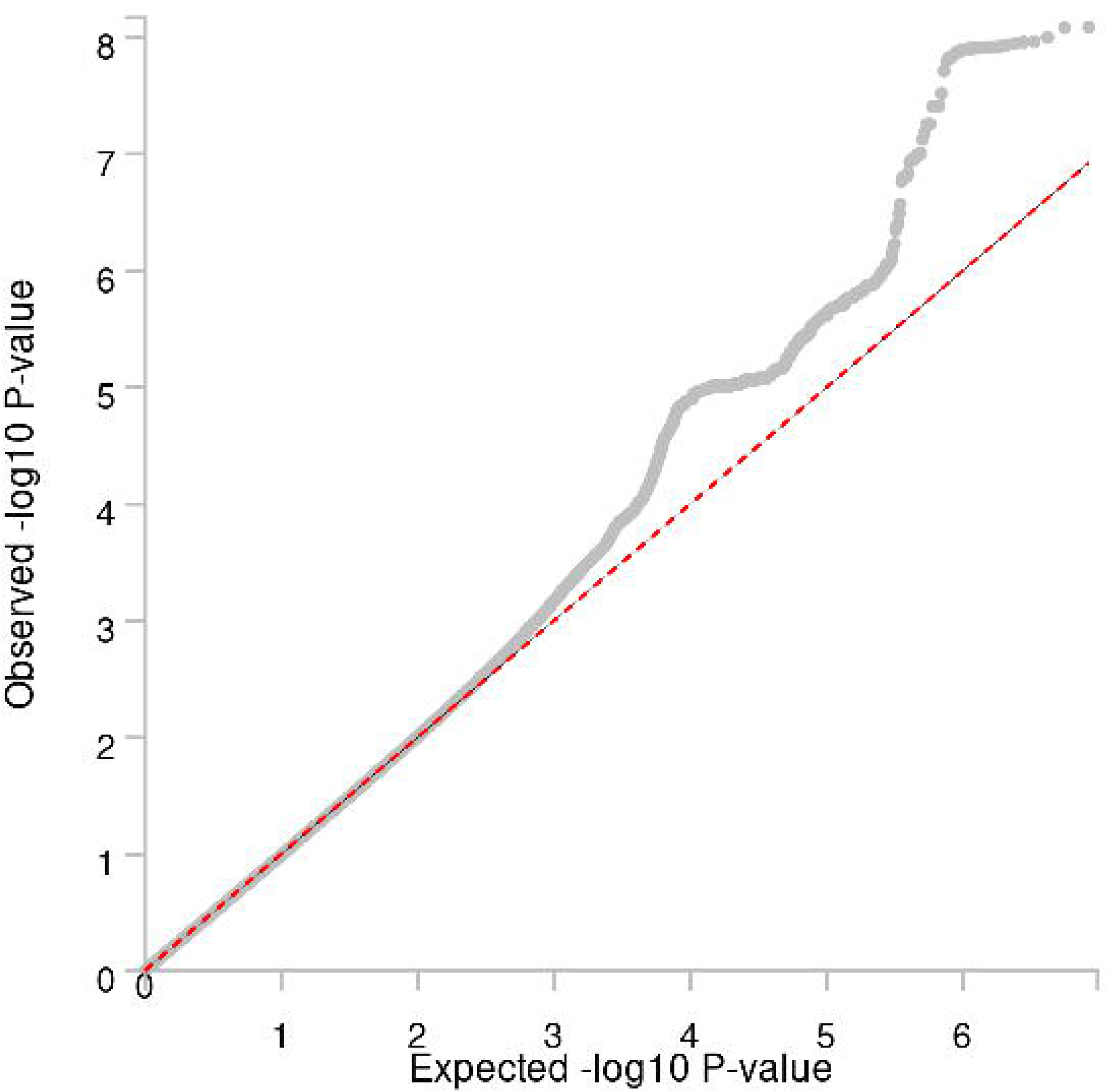

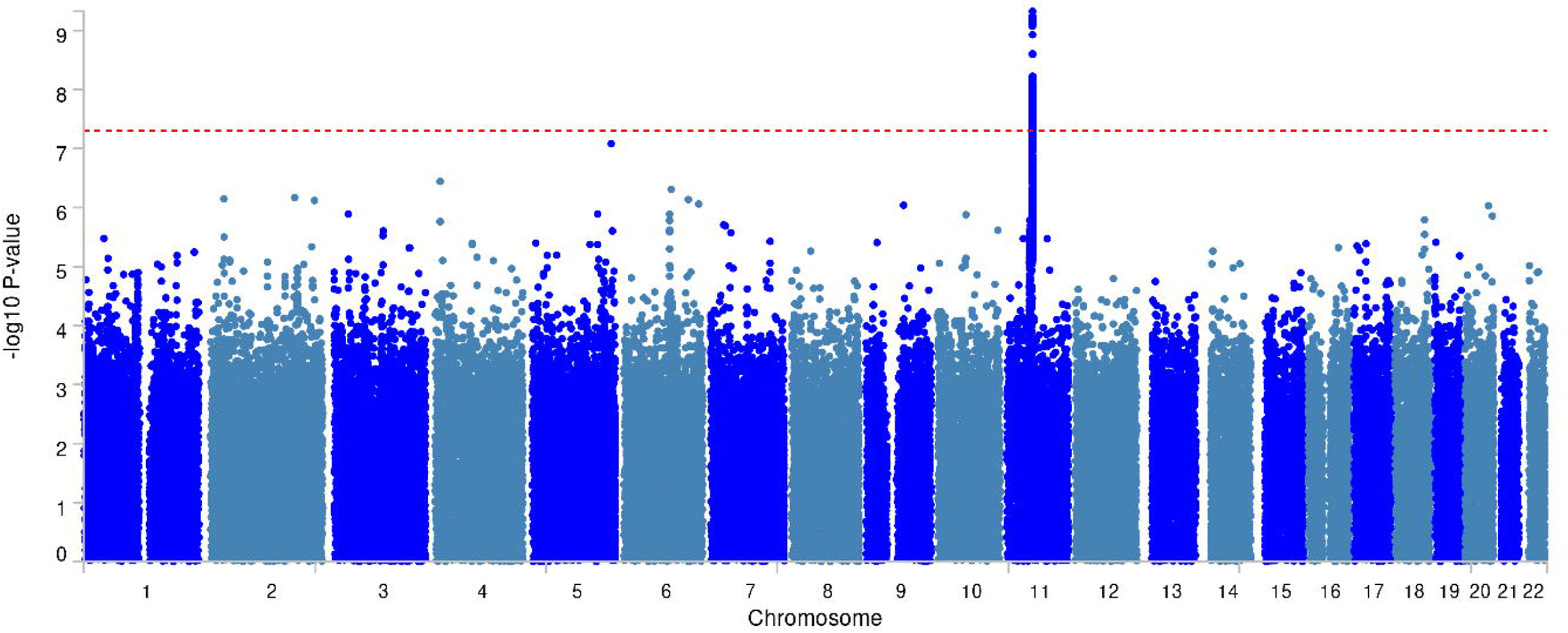

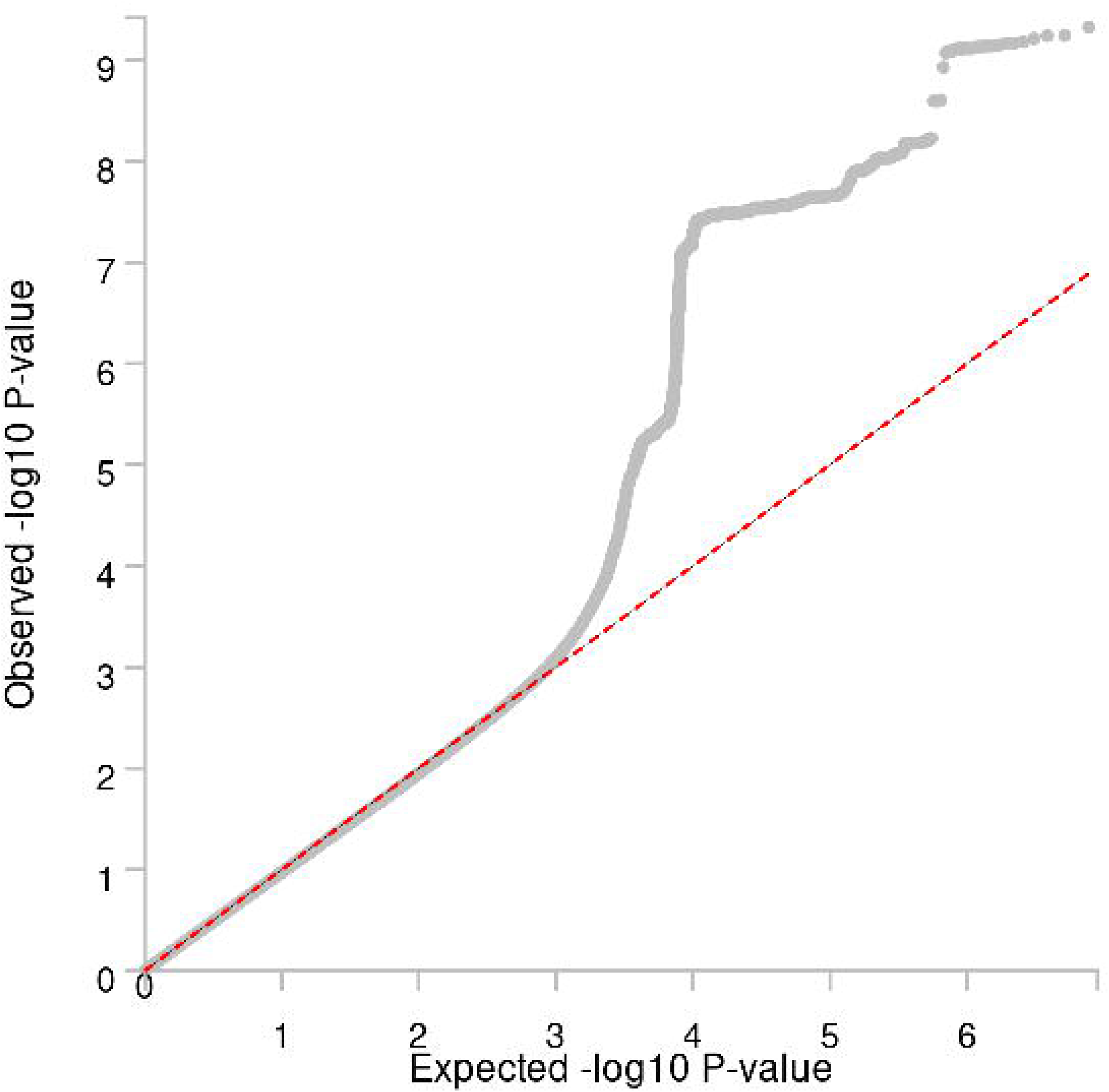

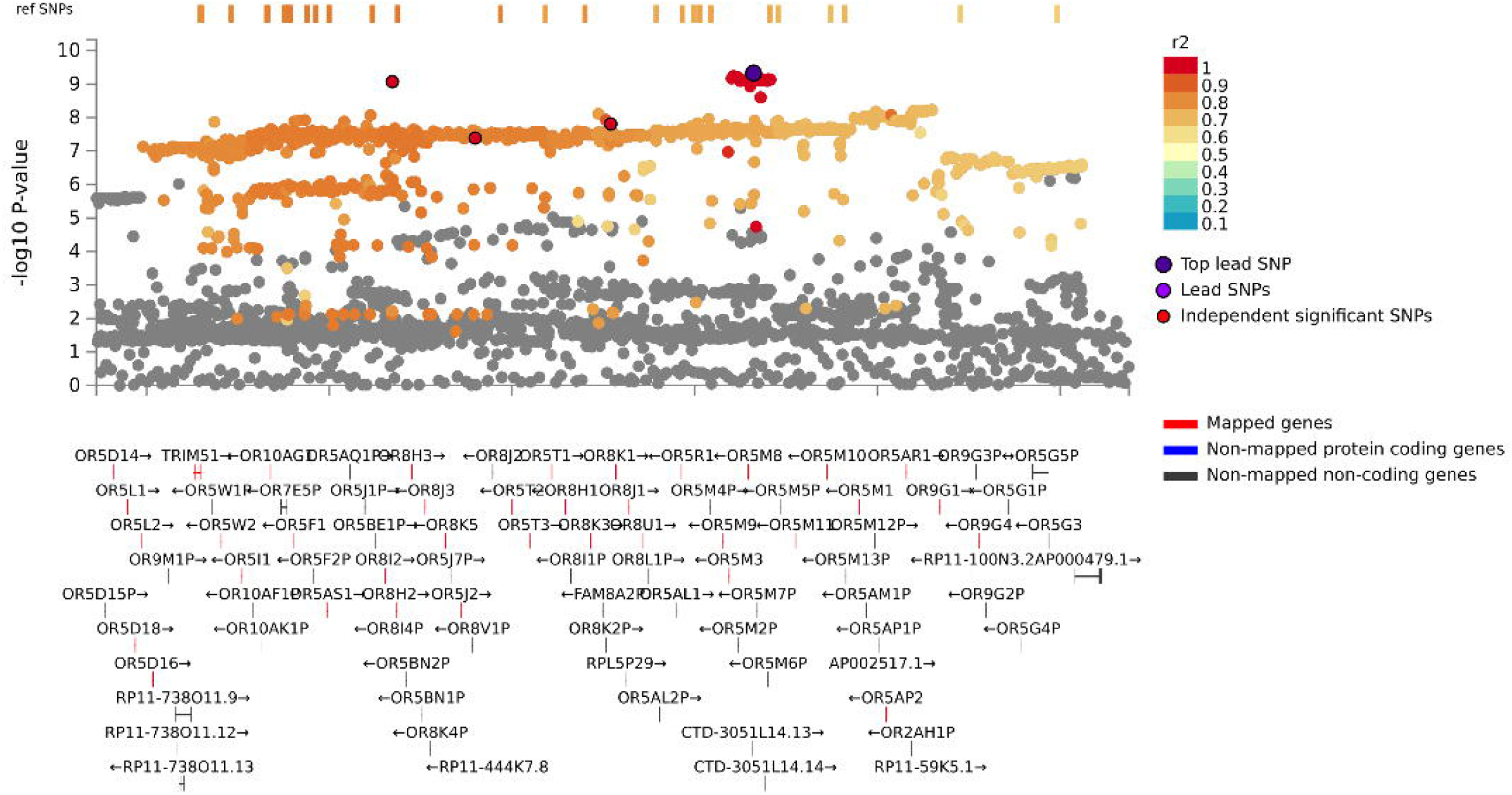

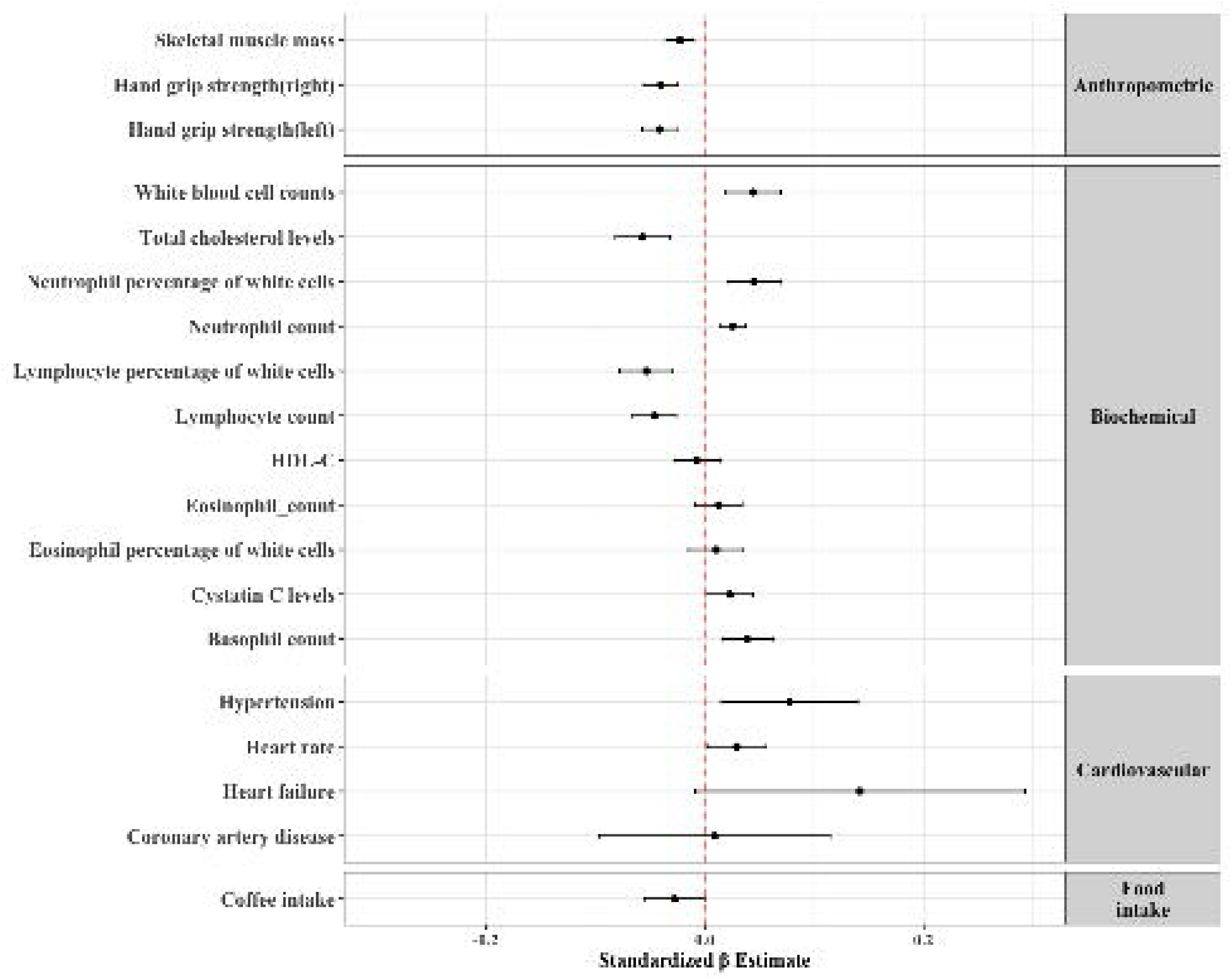

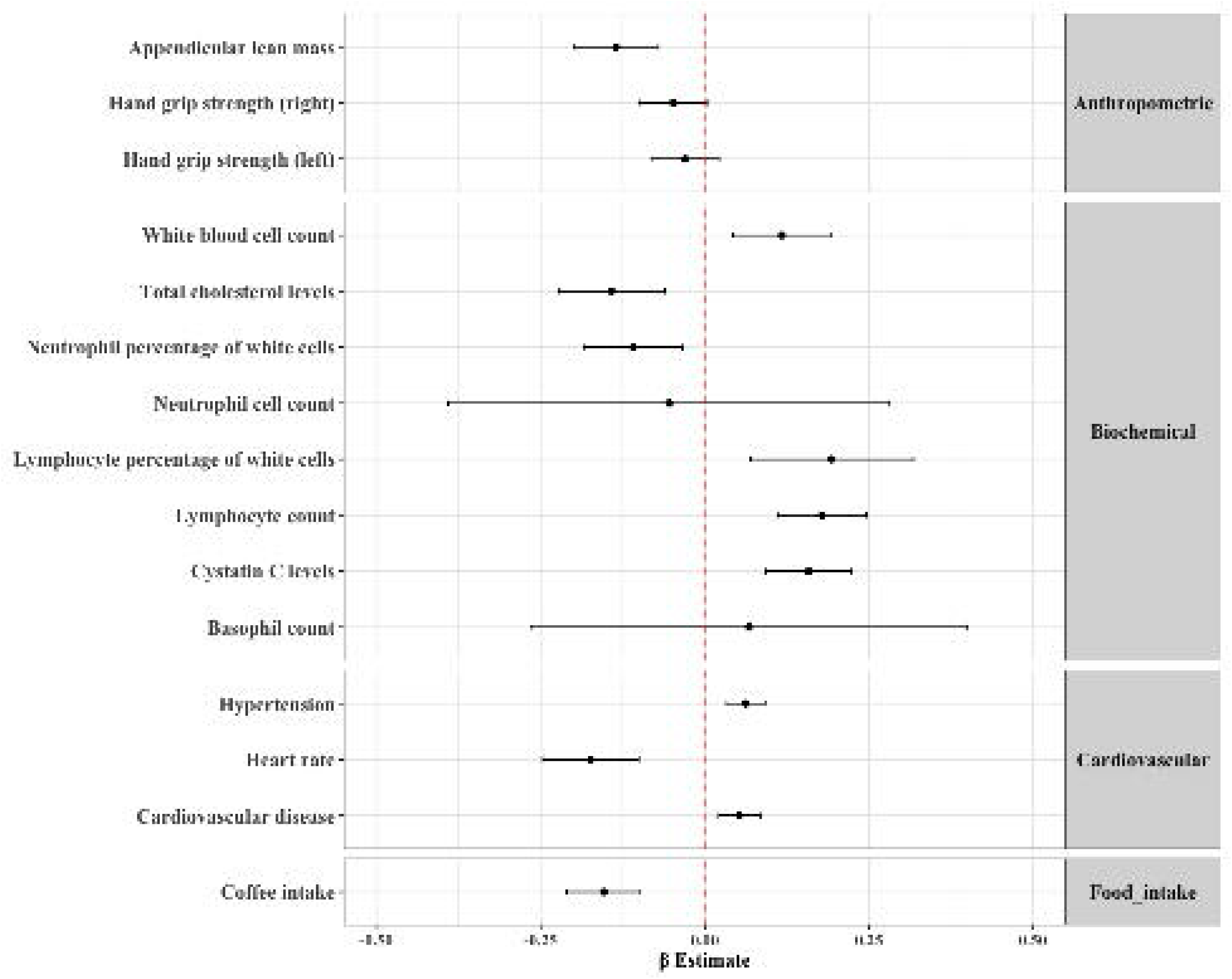

