## Supplementary Methods & Figures for "Genome-Wide Association Study Meta-Analysis Uncovers Novel Genetic Variants Associated with Olfactory Dysfunction"

**eMethods**

**eFigure 1:** Quantile-quantile plots per cohort

**eFigure 2:** Sensitivity analysis using adjusted p-values

**eFigure 3:** 3D-chromatin interaction (Hi-C) mapping

**eFigure 4:** Gene-enrichment analysis

**eTable 1:** Genomic risk loci associated with olfactory dysfunction in model 1 and model 2

**eTable 2:** Lead SNPs associated with olfactory dysfunction in model 2

**eTable 3:** Independent SNPs associated with olfactory dysfunction in model 2

**eTable 4:** Genome-wide significant SNPs that are in LD (r2>0.6) with the independent SNPs in model 2

**eTable 5:** ANNOVAR Annotation of the lead SNPs and SNPs that are in LD with lead SNPs in model 2

**eTable 6:** Genes mapped based on positional, Eqtl and chromatin interaction mapping in model 2

**eTable 7:** Chromatin interaction analysis of mapped genes in model 2

**eTable 8:** MAGMA Gene-based analysis in model 2

**eTable 9:** MAGMA Gene set analysis in model 2

**eTable 10:** Functional validation of mapped genes using gene expression analysis

**eTable 11:** PheWAS lookup of lead SNPs

**eTable 12:** Associations between olfactory dysfunction and other phenotypes using individual-level data from the Rhineland Study

**eTable 13:** Two-sample Mendelian Randomization analyses of the effect of olfactory dysfunction on other phenotypes

**eMethods**

**Cohort descriptions**

**Rhineland Study**

The Rhineland Study is an ongoing prospective cohort study enrolling individuals aged 30 or above from two geographically defined areas in Bonn, Germany. The only exclusion criterium is insufficient command of the German language required for providing informed consent. All participants underwent deep phenotyping to derive genetic, imaging, cognitive, socio-demographic as well as sense of smell information.

**ARIC**

The ARIC Study is an ongoing longitudinal study that was established in 1987–1989 to investigate risk factors for cardiovascular diseases.

**LIFE-Adult**

The LIFE-Adult-Study is a population-based cohort study investigating the prevalence and incidence of common diseases and subclinical disease phenotypes, the complex interactions between genetic and lifestyle factors regarding the co-occurrence and development of subclinical phenotypes and diseases, and the role of biomarkers to predict disease initiation and progression. The study comprises an age- and sex-stratified random sample of 10,000 adult individuals (aged 18-79 years) from Leipzig, Germany. The assessment program comprises physical and medical examinations, personal interviews, self-administered questionnaires, psychometric tests, and clinical chemistry from blood and urine samples (including biobank asservation).

**CHRIS**

The Cooperative Health Research in South Tyrol (CHRIS) Study is an ongoing longitudinal study established in 2003 to investigate genetic and lifestyle determinants of health and healthy aging in a single administrative district of the alpine Bolzano-South Tyrol province of Italy with a stable population, cooperative administration, shared single reference hospital, relatively homogenous customs and environment, and high healthy life expectancy. 13 393 adults aged ≥18 years (median 46, range 18-94, 54% female) participated in a baseline visit at the district reference hospital in 2011-2018, providing socio-demographic, health, lifestyle, and exposure data from self-report questionnaires, interviews, and instrumental examinations, plus urine and blood samples for biobanking, DNA extraction, and molecular characterization (genome and exome sequencing, metabolomics, proteomics). 3 500 baseline participants have completed a visit as part of ongoing follow-up, providing most of the same data and samples as at baseline; additional data collection from wearable activity trackers and on prominent themes like COVID-19 have been introduced.

**Olfactory dysfunction assessment**

In the Rhineland Study, the ARIC study and the LIFE-Adult study, olfactory dysfunction was assessed using the 12-item “Sniffin’ Sticks” odor identification test (SIT-12). This is assessed by using twelve felt-tip sticks from a test kit (Burghart Messtechnik GmbH, Germany), each carrying a distinct odorant, were consecutively positioned approximately 2 cm in front of both nostrils for 3 to 4 seconds by trained technicians in a well-ventilated room. Participants were then asked to choose only one out of four answer options for each odorant. The time interval between two consecutive odor presentations was at least 20 seconds. The olfactory dysfunction SIT-12 score was generated as the total number of incorrect answers (range 0-12). The CHRIS study employed the 16-item “Sniffin’ Sticks” odor identification kit for assessment of olfactory function. For the purposes of this study, results from four pens *(turpentine*, *garlic*, *apple*, and *anise*) were excluded.

**Genotyping, Quality Control and Imputation**

In the ARIC study, genotyping was performed using the Affymetrix GeneChip SNP Array 6.0, applying standardized quality-control filters for call rate (<95%), Hardy Weinberg equilibrium (HWE) (p < 1 x 10^-5) for SNPs and for the samples to exclude individuals with call rate (<95%) before imputation. In the Rhineland Study, blood samples were genotyped using the Illumina Omni-2.5 exome array and processed with GenomeStudio (version 2.0.5). Quality control was performed using PLINK (version 1.9). SNPs were excluded based on poor genotyping rate (< 99%) or HWE (p < 1 x 10^−6). Additionally, participants with poor quality DNA samples were excluded, because of poor call rate (<95%) (n=51), abnormal heterozygosity (n=100), cryptic relatedness (n=472), or sex mismatch (n=43). To account for variation in the population structure, which may otherwise cause systematic differences in allele frequencies,^1^ we used EIGENSTRAT (version 16000). EIGENSTRAT uses principal component analysis to detect and correct for population structure, which resulted in the exclusion of an additional 164 participants from non-European descent. Finally, imputation was performed using IMPUTE (version 2)^2^ and the 1000 Genomes version 3 phase 5 as the reference panel.^3^ In the LIFE-Adult study, the Affymetrix AXIOM-CEU 1 array was used for assessment of genotypes. Exclusion was based on low sample call rate (<97%), mismatches of reported and genotyped sex, duplicated samples or samples with unresolved relatedness and ethnic outliers. Imputation was performed with IMPUTE2 (Ver. 2.3.2) after prephasing with SHAPEIT (v2.r837). For the imputation the following filter criteria were used: SNP call rate < 97%, HWE < 1 x 10^−6, plate association < 1 x 10^−7, monomorphic SNPs; SNPs violating criteria of Affymetrix cluster measures FLD, HetSO and HomRO were removed too. In the CHRIS study, genotyping was carried out for consenting participants in three batches using the Illumina OmniExpressExome chip (n=5,882), the Illumina Human Omni2.5Exome chip (n=4,887), and the Illumina-based OmniEURHD chip from Life & Brain GmbH, Bonn (n=2,694). Initial processing and quality control (QC) of returned raw genotyping data was performed on each genotype batch independently at the time of acquisition using the Illumina GenomeStudio software package with additional QC following established protocols similar to those described above. Following quality control, batches were merged together consecutively, discarding variants not present on all array chips, and excluding samples with >5% missingness. The genotype dataset consists of approximately 579,000 variants from autosomal chromosomes in 12,834 samples. TOPMed imputation was carried out with the topmed-freeze5 panel^4^ on the Michigan Imputation Server (1.7.1),^5^ using minimac4-1.0.2.^6^ Excluding markers with imputation R2 < 0.3, the CHRIS TOPMed-imputed dataset contains 35,061,390 variants.

**Variant annotation and gene mapping**

Based on the standard 10 kb gap between SNPs and genes, each lead SNP was individually mapped to the gene. Variant annotation for each locus was based on lead SNPs and candidate SNPs, defined as those SNPs in LD with the lead SNP within a window of 250 kb and nominally significant at p < 0.05. Functionally annotated SNPs were subsequently mapped to genes based on (i) physical position on the genome (positional mapping), (ii) expression quantitative trait loci (eQTL) associations (eQTL mapping), and (iii) 3D chromatin interactions (Hi-C).

**Gene and Gene-set analysis**

Gene-based analyses were performed using MAGMA (**generalized gene-set analysis of GWAS)** v1.6 as implemented in FUMA, using the Bonferroni method for multiple testing. Mapped genes from SNP2GENE were further investigated using the GENE2FUNC tool in FUMA, which through a hypergeometric test assesses pathway enrichment of mapped genes in the Molecular Signatures Database (MSigDB) gene sets.

**Gene expression sequencing and analysis**

Briefly, total RNA sequencing was performed using the TruSeq stranded total RNA kit (Illumina) on a NovaSeq6000 instrument (Illumina). Genes with overall mean expression levels greater than 15 reads and expressed in at least 95% of the participants were considered for further analysis.

**RNA sequencing and gene expression analysis in the Rhineland Study**

Blood samples were collected between 7:00 to 9:45 in the morning from an antecubital or dorsal hand vein. For RNA sequencing, samples were stored in PAXgene Blood RNA tubes (PreAnalytix/Qiagen). Paxgene Blood RNA Tubes were thawed and incubated at room temperature to increase RNA yields. Total RNA was isolated according to manufactures’ instructions using PAXgene Blood miRNA Kit and the automated purification protocol (PreAnalytix/Qiagen). Differential blood cell counts (erythrocytes, neutrophils, eosinophils, basophils, lymphocytes, monocytes and platelets) were performed at the Central Laboratory of the University Hospital in Bonn, using EDTA-whole blood samples on a hematological analyzer Sysmex XN9000. RNA integrity and quantity were evaluated using the tapestation RNA assay on a tapestation4200 instrument (both from Agilent). We used 750 ng of total RNA to generate NGS libraries for total-RNA sequencing using the TruSeq stranded total RNA kit (Illumina) following manufacturer's instructions with Ribo-Zero Globin reduction. We checked library size distribution via tapestation using D1000 on a Tapestation4200 instrument (Agilent) and quantified the libraries via Qubit HS dsDNA assay (Invitrogen). We clustered the libraries at 250 pM final clustering concentration on a NovaSeq6000 instrument using S2 v1 chemistry (Illumina) in XP mode and sequenced paired-end 2*50 cycles before demultiplexing using bcl2fastq2 v2.20. Quality control of the sequencing was evaluated through FastQC v0.11.9. Following the trimming of low-quality score reads through Trimmomatic v.0.39 software, sequencing reads were aligned using STAR v2.7.1 and the human reference genome GRCh38.p13 provided by Ensembl. The count matrix was generated with STAR–quantMode GeneCounts using the human gene annotation version GRCh38.101. Genes with overall mean expression greater than 15 reads and expressed in at least 95% of the participants were included for further analysis. Raw counts were normalized and transformed using the varianceStabilizingTransformation function from DESeq2 (v1.30.1). We extracted the expression levels of mapped genes from total RNA sequencing data available from a subset of Rhineland Study participants (n=1985). In brief, multivariable linear regression was used to assess the association between z-transformed gene expression levels and olfactory dysfunction, while adjusting for age, sex, APOE*ε4 carrier status, the first 10 genetic principal components, global cognitive function and sequencing batch. Subsequently, we tested whether this association was modified by age by including an interaction term between age and gene expression levels. For genes with a significant age interaction, we performed the linear regression analysis after stratifying in age terciles (30 to 50, 50-62 and 62-95 years).

For SNPs identified in the GWAS meta-analysis and their mapped genes, functional validation of the association between SNP and gene expression was performed using multivariable linear regression. Gene expression was coded as the dependent variable and the number of effect alleles (0, 1 or 2) was coded as a numeric independent variable, controlling for age, sex, the 10 first genetic PCs and sequencing batch.

**Phenome-wide association studies**

We used the Open Target Genetics platform ^7^ to perform Phenome-Wide Association Studies (PheWASs). This platform searches for the association of the lead variant across various phenotypes/diseases present in largescale GWAS databases such as the GWAS catalogues of the National Human Genome Research Institute – European Bioinformatics Institute (NHGRI-EBI) and the UK biobank. The Benjamini-Hochberg false discovery rate (FDR) method was used for multiple comparisons correction.

**Demographic and phenotypic variables in the Rhineland Study**

Age, sex and smoking data were based on self-reports using questionnaires. Smoking was coded as a dichotomous variable (current *vs.* non-current smoker). Participants with missing data on smoking were classified as current smokers when their cotinine metabolic levels, measured with the Metabolon HD4 platform, exceeded the non-smoker sample-defined 97.5 percentile. Body mass index (BMI) was measured as weight (kg) divided by height squared (m^2^). Skeletal muscle mass (kg) was derived from bioimpedance analysis. Muscular strength was measured using the hand-held Jamar Plus Digital Dynamometer (Patterson Medical, USA). The grip force of each hand was measured three times and the average of the hand grip strength was calculated. Hypertension was coded as “yes” in case of antihypertensive drug use or high blood pressure (mean systolic blood pressure >=140 mmHg or diastolic blood pressure >=90 mmHg), and as “no” otherwise. Heart rate was measured as number of heart beats per minute. Cardiovascular conditions including stroke, heart failure, and coronary artery disease (CAD) were defined as self-reported physician diagnosis. Differential blood cell counts (e.g., erythrocytes, leukocytes, basophils, eosinophils, lymphocytes, monocytes, neutrophils) were measured at the Central Laboratory of the University Hospital in Bonn using EDTA-whole blood samples on a hematological analyzer Sysmex XN9000. Habitual dietary intake (ml/day) was assessed by a self-administered semi-quantitative food frequency questionnaire (FFQ). We assessed whether phenotypes associated with genetic variants of olfactory dysfunction (after FDR correction) were also associated with a poor sense of smell on a phenotypic level using multivariable regression models. Statistical significance was inferred at p < 0.05. We log-transformed white blood cell (WBC), eosinophil, neutrophil and lymphocyte cell counts, as well as skeletal muscle mass, to account for their skewed distributions. All the phenotypes were adjusted for poor sense of smell and further corrected for age and sex. We additionally adjusted for potential risk factors such as smoking when the dependent variable was heart rate, white blood cell counts for neutrophil, eosinophil and lymphocyte cell counts and BMI for skeletal muscle mass and cardiovascular diseases (CVD). All the numerical variables were standardized to a mean of 0 and a standard deviation of 1 to allow for better comparison of the effect sizes across different traits.

**Two-sample Mendelian Randomization (MR)**

For Two-sample MR, we included those phenotypes as outcomes that were identified in the PheWAS analysis and were also significantly associated with olfactory dysfunction on a phenotypic level in the Rhineland Study population. As genetic instruments for the exposure we used genome-wide significant SNPs from the olfactory dysfunction GWAS meta-analysis. SNPs in LD were clumped based on r^2^ < 0.01 at a 10 Mb window before the MR analyses.

**eFigure 1. Quantile-quantile plots per cohort.** Quantile-Quantile plots for the GWAS-summary statistics on olfactory dysfunction in people of European ancestry in each cohort in model 2.

A) **RHINELAND STUDY	B)	ARIC**

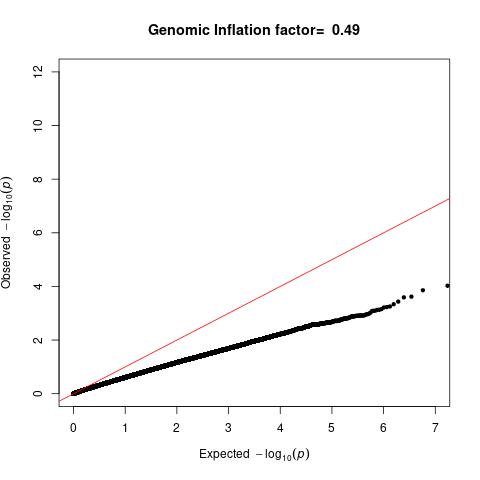

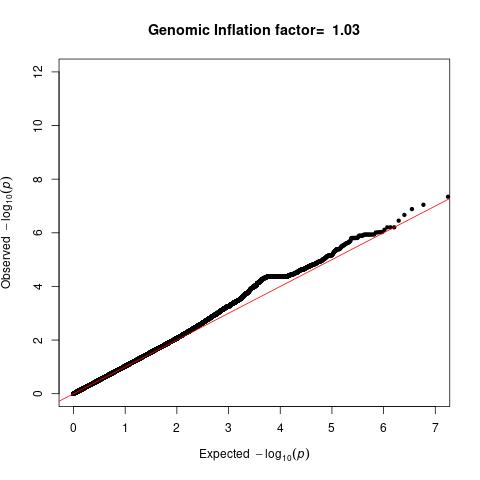

**C)	LIFE-Adult	D)	CHRIS**

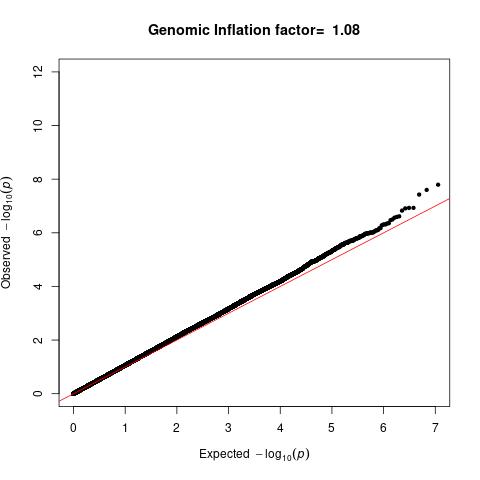

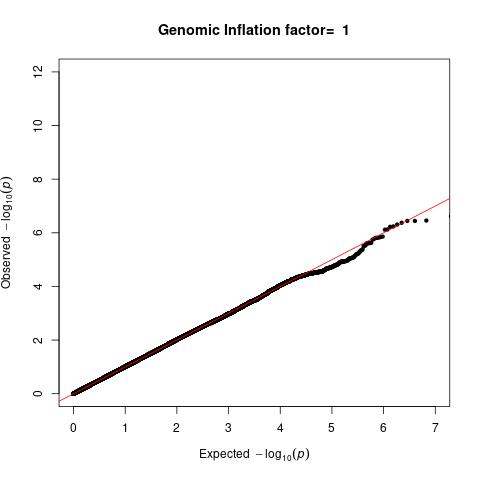

**eFigure 2. Sensitivity analysis using adjusted p-values.** Quantile-Quantile plot for the GWAS-summary statistics from model 2 in ARIC cohort using adjusted p-values (**A**). Manhattan plot (**B**) and corresponding quantile-quantile plot (**C**) of the genome-wide meta-analysis in the European ancestry participants after inclusion of the adjusted p-value summary statistics from the ARIC cohort. The horizontal red dashed line indicates the threshold for genome-wide significance (p < 5 x 10^-8)

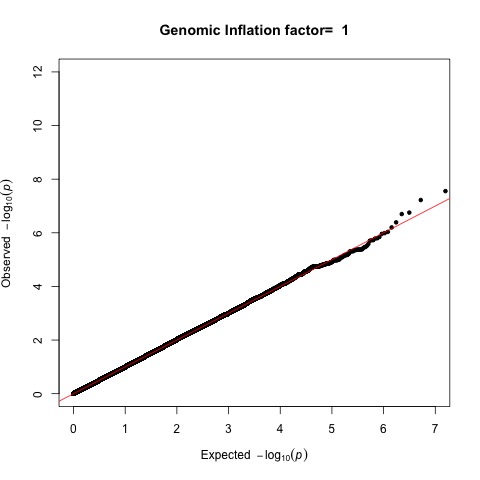
**A)**

**B)	C)**

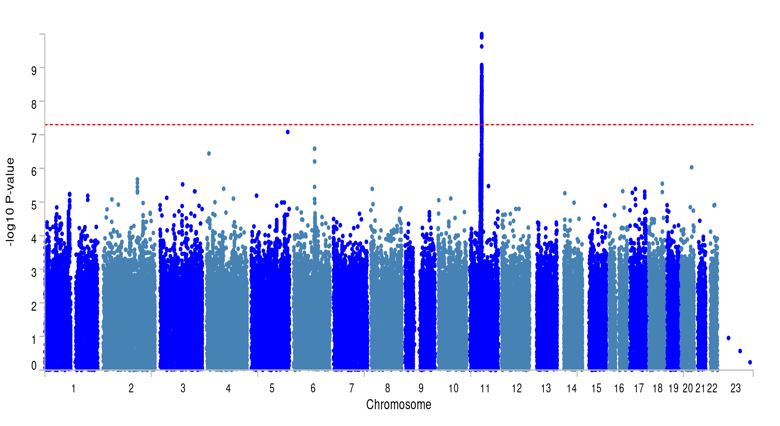

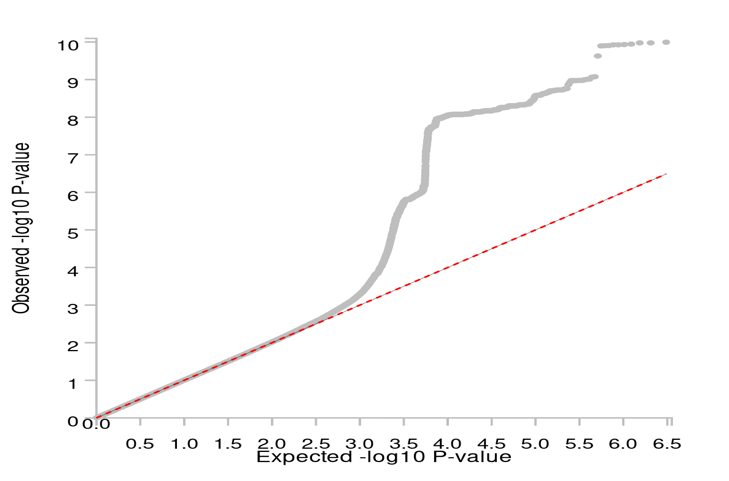

**eFigure 3. 3D-chromatin interaction (Hi-C) mapping**. Hi-C revealed significant interactions between genetic variants in *OR5M8* and other genes on chromosome 11 (FDR < 1 x 10^-6), shown in orange.

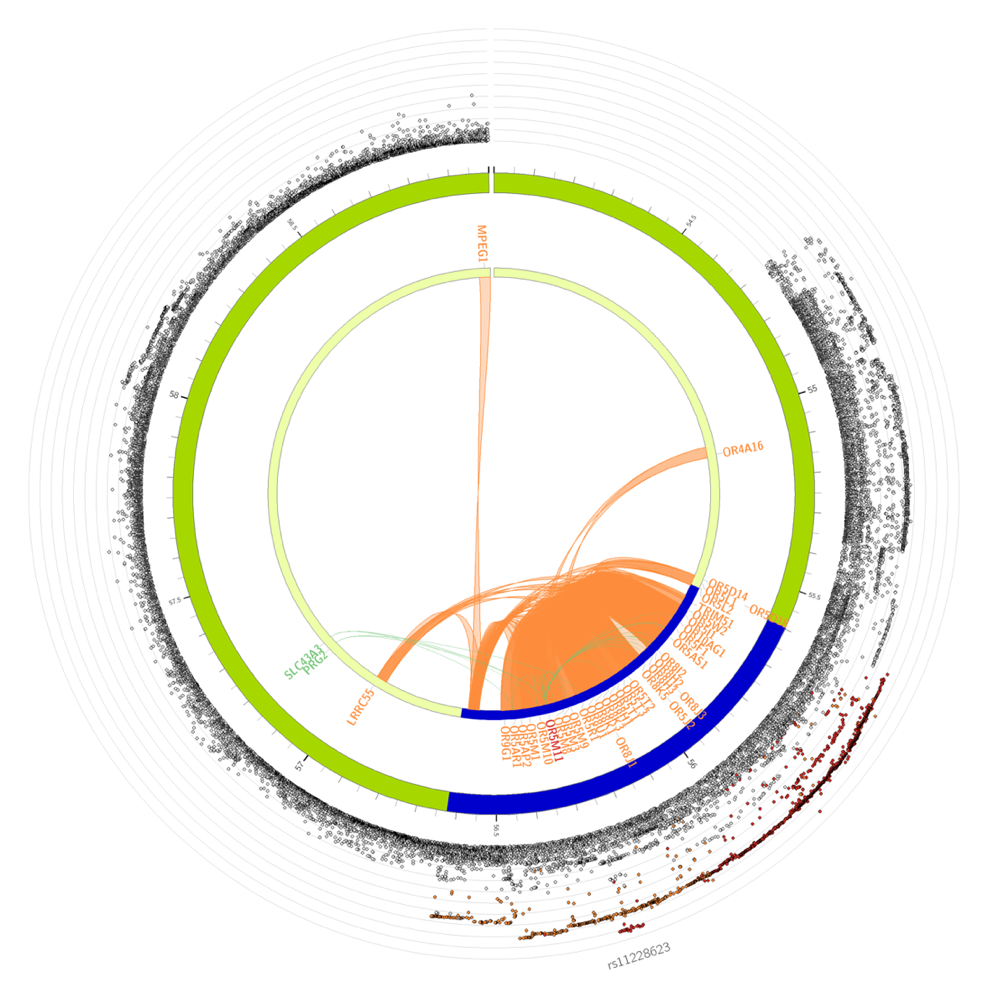

**eFigure 4. Gene-enrichment analysis.** Enrichment analysis of mapped genes, according to Gene ontology (GO): biological process (**A**), GO: molecular functions (**B**), and Kyoto Encyclopedia of Genes and Genomes (KEGG) (**C**). Pathways and processes that are overrepresented in the gene set of interest are shown. Input genes that are overlapping in the pathway or process, the enrichment p−value and the proportion of the overlapping genes (input genes relative to the tested gene set) are shown.

**A)**

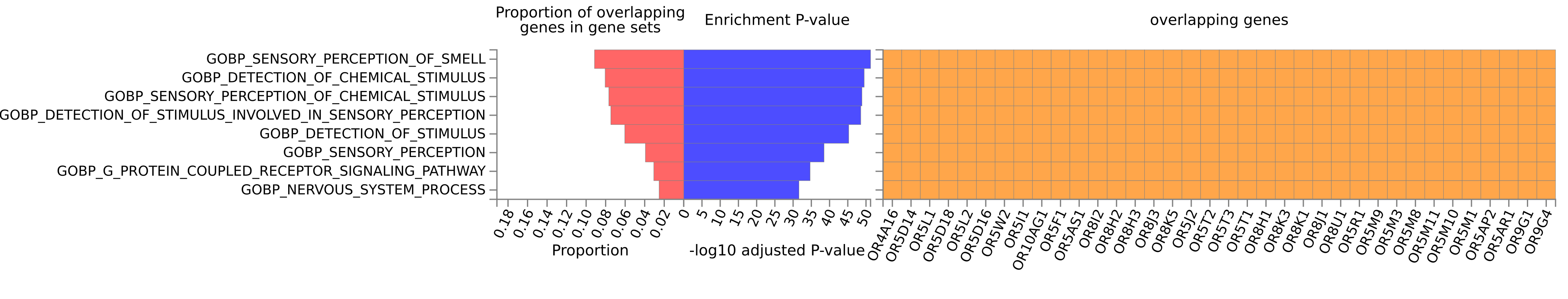

**B)**

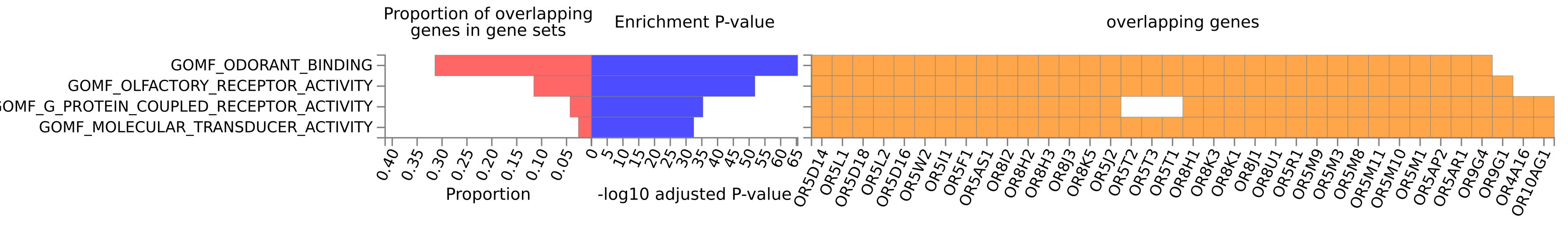

**C)**

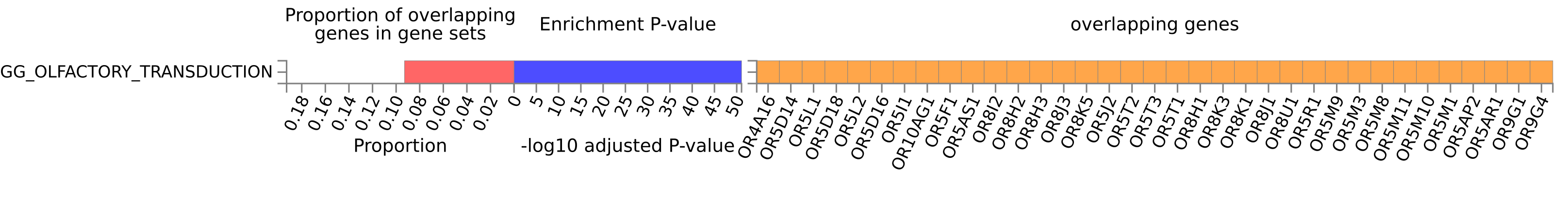
